# Graphene Field Effect Biosensor for Concurrent and Specific Detection of SARS-CoV-2 and Influenza

**DOI:** 10.1101/2022.10.04.22280705

**Authors:** Neelotpala Kumar, Dalton Towers, Samantha Myers, Cooper Galvin, Dmitry Kireev, Andrew D. Ellington, Deji Akinwande

## Abstract

The SARS-CoV-2 pandemic has highlighted the need for devices capable of carrying out rapid differential detection of viruses that may manifest similar physiological symptoms yet demand tailored treatment plans. Seasonal influenza may be exacerbated by COVID-19 infections, increasing the burden on healthcare systems. In this work, we demonstrate a technology, based on liquid-gated graphene field-effect transistors, for rapid and ultraprecise detection and differentiation of influenza and SARS-CoV-2 surface protein. Most distinctively, our device consists of 4 onboard graphene field-effect electrolyte-gated transistors arranged in a quadruple architecture, where each quarter is functionalized individually (with either antibodies or chemically passivated control) but measured collectively. Our sensor platform was tested against a range of concentrations of viral surface proteins from both viruses with the lowest tested and detected concentration at ∼50 ag/mL, or 88 zM for COVID-19 and 227 zM for Flu, which is 5-fold lower than the values reported previously on a similar platform. Unlike the classic Real-Time Polymerase Chain Reaction (RT-PCR) test, which has a turnaround time of a few hours, our technology presents an ultrafast response time of ∼10 seconds even in complex media such as saliva. Thus, we have developed a multi-analyte, highly sensitive, and fault-tolerant technology for rapid diagnostic of contemporary, emerging, and future pandemics.

## Introduction

The pathology of upper respiratory viruses has regularly presented challenges to global health care systems and their resources. The emergence of new virus variants that can evade communal immunological memory can be rapidly transmitted through airborne mucosal droplets, often resulting in the emergence of sudden seasonal epidemics or pandemics. Over the last century, the most prominent of these viruses have been variants of influenza (Flu), which have been estimated to be responsible for approximately 400,000 deaths annually^1^. The emergence of the novel coronavirus SARS-CoV-2 (COVID-19) in 2019 introduced a new upper respiratory virus that, as of now (April 2023) has led to at least 6.8 million deaths globally ^2^.

The COVID-19 pandemic has highlighted the need for new rapid point of care diagnostic systems for upper respiratory viruses, especially for high population density areas where the transmission can be the most potent and diagnostic availability and turnaround time the most limited. Significant challenges in respiratory diagnostics include the establishment of assays with a limit of detection (LoD) suitable for identifying early infections, minimizing false positive rates, and reducing the time to perform the assay. The current standard, the reverse transcription polymerase chain reaction (RT-PCR) isn’t ideal for identifying early respiratory infections, as demonstrated by the United States’ Center of Disease Control’s (CDC) recommendation that these assays should be performed 5 days after an exposure to ensure maximal viral titer^3^. Additionally, RT-PCR assays typically take a few hours to perform and often require transporting samples to professional laboratories, which can take a few additional days thus being a challenge during periods of high demand.

COVID-19 and Flu exhibit similar physiological symptoms ^4, 5^ underscoring the requirement for a rapid diagnostic tool capable of differentially diagnosing COVID-19 and Flu. An initial assessment of the potential cause of illness would allow a timely personalized treatment plan for the patient, thus not only aiding in curbing the spread, but also in utilizing medical resources in an efficient manner. As the recent COVID-19 pandemic spurred the rapid development of multiple COVID-19 detection platforms involving nanotechnology mediated biological tests^6^ that serve as an alternative to RT-PCR and electrical tests with varying degrees of usability and success. Although highly accurate with the ability to provide data to accurately determine the infection status post COVID-19, these tools are extremely labor intensive with slower turn-around time. To address the gap posed by the standard biology tests antibody-modified graphene field effect transistors (GFETs) have stood out due to their low LoDs and fast response time^7–14^. Prior to COVID-19, GFETs had already demonstrated their exceptional capability through Flu diagnostics platform^13, 15^. Imbibing these GFETs with concurrent multiple target detection capability would increase their effectiveness not only during pandemics but also in instances where there is an urgent requirement to detect the cause of illness in a patient showing overlapping symptoms with another disease.

In this work, we have developed a concurrent rapid differential diagnosis platform using antibody-modified GFET. The device is a holistic platform having 4 onboard GFETs isolated from each other using polydimethylsiloxane (PDMS) barriers yet enclosed in a higher perimeter PDMS wall (Figure 1a and Figure S1) so that they can be functionalized individually in isolation and tested using a shared biological sample without the assistance of complex microfluidics. The enclosure also assists in conducting multiplexed detection of COVID-19 and Flu without the deployment of microfluidics. Each GFET is modified with either an antibody of interest, i.e., COVID-19 or Flu or are used as a control. The device design enables isolated targeted functionalization of graphene channels while allowing a common medium for introducing the analyte, which then translates into common gating and a change in conductance of the GFET modified with the corresponding target/receptor^16^. In this case, the chip has two GFETs dedicated to antibody immobilization for COVID-19 and Flu each, while one GFET was only chemically passivated with Tween-20 (Tw20) and another left bare as a control (Figure 1b). The presence of onboard control is an excellent example of an integrated control on FET devices which has not been demonstrated in any other graphene FET based work in our knowledge. This makes our device very close to the commercially available immunoassay devices that also come with integrated control and don’t need to be separately tested against other samples. This proof-of-concept device can be translated towards manufacturing which would substantially bring down the costs. As has been demonstrated by GFET based rapid diagnostics company^17^, these types of devices can be operated with a handheld reader designed to read out and interpret the electrical results.

**Figure 1.**
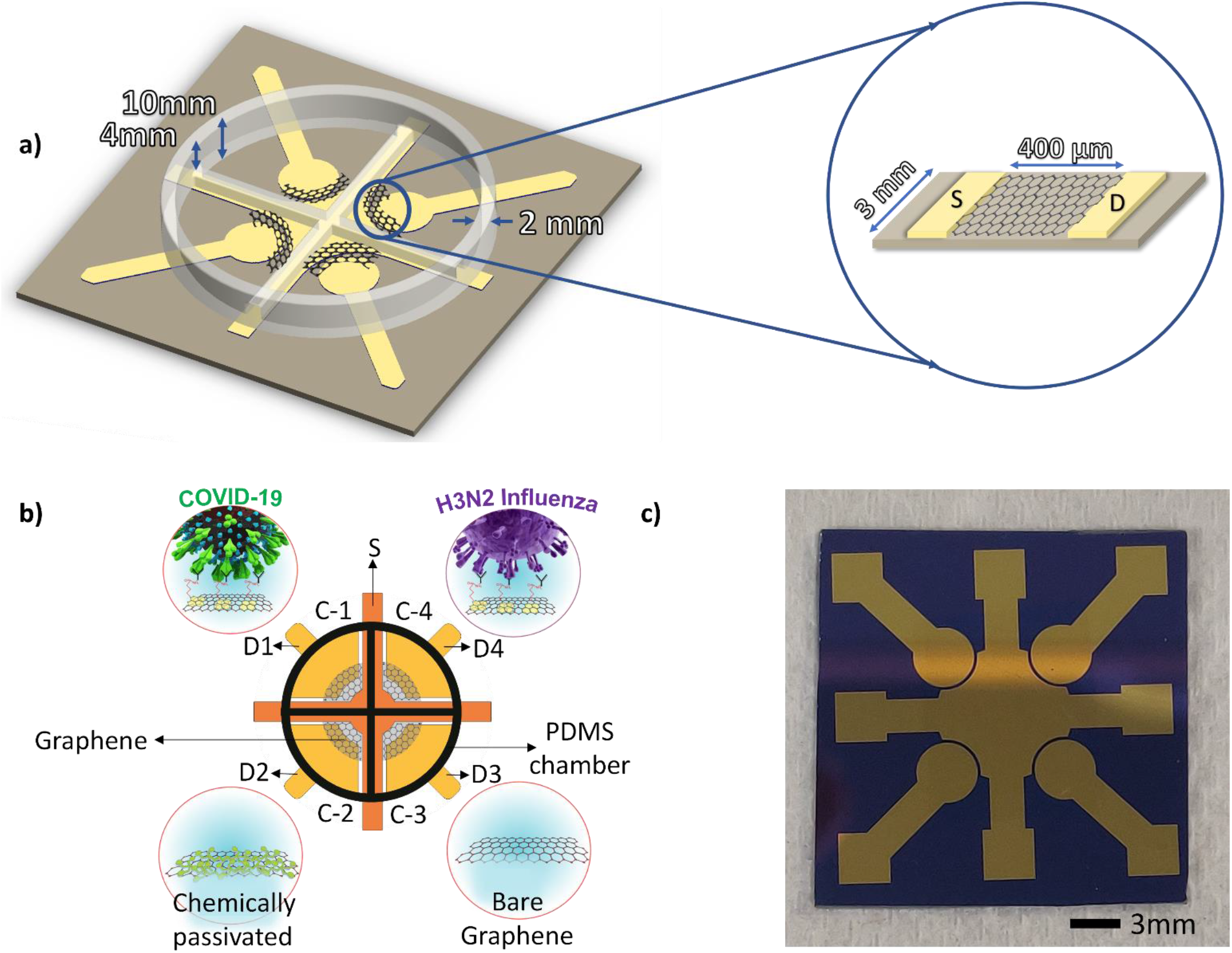
Schematic of the COVID-19 and Flu dual detection biosensor. a) Dimensions of the device, the 6mm tall inner cross of the device ensures isolated functionalization of each GFET. The length of channel in each transistor is 400µm while the width is around 3mm. Each GFET quarter can accommodate at least 25µl of fluid without any issue of interference. (b) Schematic diagram of the dual detection sensor: C-n stands for the array n-FETs in the device resulting in a four-channel arrangement. S denotes the common source electrode across all the FETs onboard. D-n, stands for specific drain electrode corresponding to each individual FET. C-1 and C-4 are immobilized with CR3022 and FI6v3 antibodies, respectively, C-2 is chemically passivated with Tw20 as a comparative control, while C-3 is bare. (The graphene channel length is 400µm with a width of 3mm) (c) Optical image of the sensor with 4 GFETs. Immobilization process flow of the GFETs.

Our antibody immobilized GFETs have registered the lowest measured concentration of the COVID-19 Spike protein and the Flu surface protein, Hemagglutinin (HA), at around 88zM and 227zM, respectively. Combined with almost negligible cross-reactivity, we can claim a fast and specific response with the reaction time of ∼10s depending on the antigen. Together, the performance of the proposed devices opens the possibility of diagnosing patient’s conditions well ahead of the 5-day gap suggested by the CDC thus helping in curbing the spread of disease.

## Results and Discussion

### Device schematic and working

Our GFETs are fabricated in a photolithography free process thus presenting a minimal chance of additional graphene contamination through lithography chemical residues. This, we posit, allows for more effective functionalization and cleaner graphene and electrode interface thus leading to higher sensitivity. Shortly, we begin with evaporation of metal electrodes through a shadow mask, followed by classic PMMA-assisted wet transfer of graphene. However, here we transfer rather large pieces of graphene (1×1 cm^2^), that covers all four quadrants (Figure 1a-b and Figure S1) at the same time. After the transfer, PMMA is etched, and the clean graphene is exposed to the environment.

Each device consists of an array of 4 GFETs presenting 4 channels of operation (C-n), isolated from each other through PDMS enclosures (Figure 1b). The ratio of the height of the inner enclosure in the form of a cross with respect to the outer enclosure has been set at 0.6 (Figure 1a and Figure S1), where the inner enclosure is shorter than the outer enclosure. The height difference between the inner and outer enclosure allows for independent functionalization of each GFET while allowing all the GFETs to be driven through a common gate operating with a common medium during measurements. The length of channel in each transistor is 400 µm while the width is around 3mm. Each GFET quarter can accommodate at least 25 µl of fluid without any issue of interference. An Ag/AgCl pellet-based electrode is submerged into the shared medium to act as the gate electrode.

### Biological Reagents

To distinguish between viruses, we selected antibodies that recognize a unique antigen for each virus. For COVID-19, we used the antibody CR3022 to target the receptor-binding domain (RBD) region on the transmembrane Spike protein. For Flu, we selected the engineered antibody FI6v3 to bind to the conserved central stalk domain of transmembrane protein hemagglutinin (HA). The antibodies selected are each capable of binding to multiple variants of their respective virus. For COVID-19 antibodies, it has been reported that those that can recognize the spike protein are often able to cross react with other variants of the virus although with different affinities^18^. For CR3022, it has been shown to be capable of recognizing most of the common variants of COVID-19^19, 20^. Additionally, FI6v3 was engineered to bind to all type 1 and 2 influenza A subtypes^21^. The diversity of variants that can be recognized gives this assay tremendous breadth among different subtypes of each virus. The interaction between the antibodies and their respective analyte proteins was validated through ELISA for each batch of antibodies (Figure S2). It is important to note here that unlike comtemporary^10, 22–25^ GFET biosensors, our devices were made in photolithography-free environment, hence featuring a relatively clean surface that enables effective functionalization with PBASE and consecutively, antibodies along with cleaner interface between graphene and electrodes. Thus, in turn, affording the ultrahigh sensitivity.

### Device working principle

The electric double layer (EDL) formed at the graphene electrolyte interface serves as a dielectric layer^26^. The common electrolyte enabling the operation of the GFETs is a low ionic strength PBS set at 0.01X. The decision to employ PBS 0.01X was to counter the charge screening effect^27, 28^ observed in high ionic concentration solutions, which reduces the observed signal strength^29^ resulting from the interaction of the target and analyte. It is imperative that EDL fall at the range suitable for IgG antibody interactions, around 4 to 14.5 nm^30^ as opposed to the low 0.7 nm above the surface EDL formed by PBS 1X ^27^. Through our experimentation, it was observed that PBS 0.01X served as the best concentration for signal detection while also maintaining bio-molecular integrity as observed through enzyme-linked immunoassay (ELISA) (Figure S2).

### Graphene functionalization and characterization results

To allow targeted detection, the GFET channels were modified through biochemical functionalization (Figure 2a), starting with making CVD-grown graphene suitable for antibody immobilization. The lack of reactive sites or dangling bonds on CVD graphene ^31, 32^ offered no site for target immobilization, which was resolved through incubation of 1-pyrenebutanoic acid succinimidyl ester (PBASE)^33, 34^ on the surface of graphene. PBASE is a pyrene-based succinimide ester that utilizes the π-π bonds extending out at the surface of graphene. The successful immobilization of PBASE on graphene was confirmed through Raman spectroscopy and electrical characterization. Figure 2b shows the occurrence of a peak at 1623 cm^-1^ after functionalization of graphene with PBASE, which is concurrent with the presence of pyrene resonance, indicating that PBASE successfully attached to the surface of graphene^31^. The reduction of I_2D_/I_G_ ratio from 2.99 to 1.219, from bare to PBASE functionalized graphene, indicates disordered surface further signaling the presence of PBASE ^31, 35^, while the rightward shift of the 2D peak by 1.3 cm^-1^ is indicative of hole doping^22^. Hole doping, being an indicator of PBASE incubation on graphene^31, 35^, was also confirmed through electrical characterization (Figure 2c-d) since the IV curves denote the movement of the charge neutrality point (CNP) rightwards relative to CNP at bare graphene. The CNP at around 0.1V in bare graphene is reflective of doping introduced due to Poly(methyl-methacrylate) (PMMA) residue during the fabrication stage (Figure 2d)^36^. The right shift of the CNP indicates the successful stacking of PBASE onto graphene^31^.

**Figure 2.**
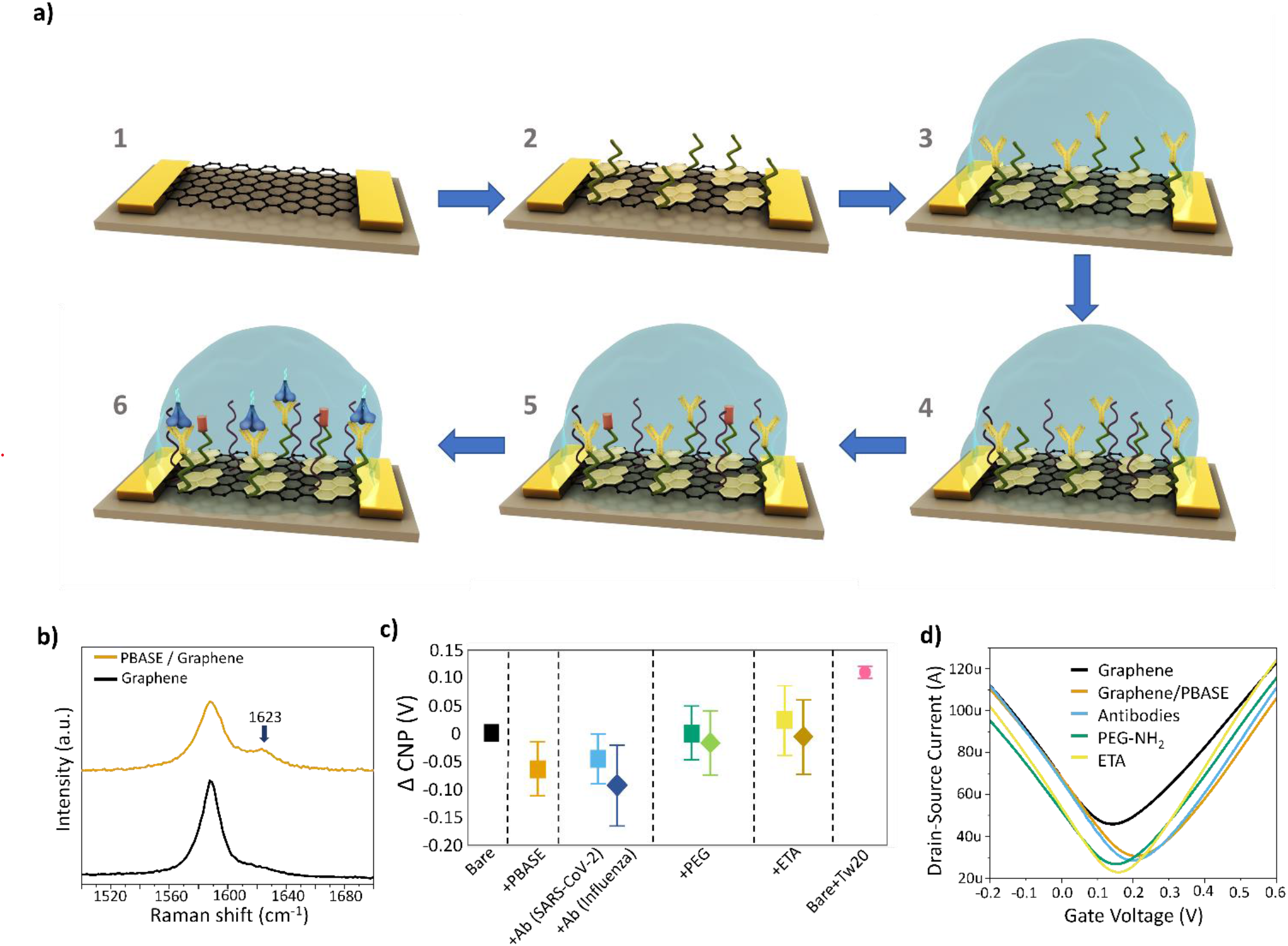
Immobilization process flow of the GFETs and characterization: (a) 1) Bare graphene transferred onto silicon via PMMA assisted wet transfer, 2) PBASE immobilization to enable linker assisted attachment of the COVID-19/Flu antibodies, 3) Immobilization of COVID-19/Flu antibodies, 4) Blocking with PEG-NH_2_ to block the unoccupied area of Graphene channel, 5) ETA assisted blocking to quench the PBASE sites that remained unoccupied by the antibodies, 6)Device operation: Interaction between COVID-19/Flu antibodies and Spike/HA proteins. (b) Raman spectroscopy of bare graphene (black) and after modification with PBASE (yellow ochre). (c) Relative change in Charge Neutrality Point (CNP) at each step of functionalization with respect to bare graphene. Whiskers are ±SD. (d) Transfer curves of the GFET after each step of functionalization (Black: bare graphene, Yellow Ochre: Graphene with PBASE; Sky blue: Antibodies, Green: PEG-NH_2_; Yellow: ETA).

The N-hydroxy succinimide (NHS) ester group in PBASE reacts with primary amine groups of the proteins, thus allowing antibody immobilization^37^. The inner cross design of the PDMS enclosure allowed specific immobilization of the CR3022 and FI6v3 onto separate GFETs on the device. The functionalization of the graphene channels was electrically monitored through the IV curves. The difference in the ΔCNP values of the CR3022 and FI6v3 as observed in Figure 2c can be attributed to isoelectric point of the monoclonal antibodies. Since CR3022 has an isoelectric point of 6.2 ^38^ it assumes a net positive charge in the measurement buffer which is PBS 1X with a pH of 7.4. The net positive charge leads to n-doping effect on the graphene channel observed in Figure 2c and Figure S5 which has also been observed by Hoang et al^39^. Similarly, a slightly negative change in ΔCNP due to FI6v3 immobilization after PBASE immobilization indicates further p-doping. This can be potentially attributed to Flu antibody’s Isoelectric point (8.4)^40^. Since functionalization takes place in 1X PBS buffer (pH 7.4), the Flu antibody assumes a net total of negative charge in turn causing p-doping effect on graphene. It should be noted that doping effects caused by the isoelectric points of the conjugated antibodies is a feature that can be modified. We have previously reported on engineering efforts to design supercharged antibodies. These are antibodies in which charged amino acids were introduced to improve the stability of the antibody while maintaining binding affinity towards the target antigen^41^.

The density of the antibody chosen for immobilization has an important role in determining the sensitivity of the device. As opposed to the expected trend that a higher density of antibodies may lead to better sensitivity, an optimal density of antibodies serves better in achieving the same. Very high density may lead to more antigen-antibody binding activity, but it will also hinder the mass transfer flux of the electrolyte towards the electrode. As such an optimal density is paramount to allow the space for mass transfer flux of the electrolyte to the electrode translating a lower density of the antibodies on the surface. This would ensure the recognition of the mass transfer flux movement of the electrolyte created due to antigen-antibody binding events^42^. In our experiments we found that the concentration of 50 µg/mL which combined with the area of our working electrode, graphene, yielded a sensitive detection range.

To ensure that the area of graphene that remained unoccupied by PBASE and the antibodies did not lead to any non-specific reaction, PEG-NH_2_ was introduced as the blocking reagent^17^. The PEG-NH_2_ plays an essential role in combatting the screening effect introduced by the electrolyte. According to previous studies^43–45^ it elevates the layer from which the screening (Debye length) occurs. Hence, the charged protein molecules can come in closer contact with the antibodies, providing a greater and faster response. To neutralize PBASE sites unoccupied by antibodies, ETA was used as the blocking agent to prevent any non-specific reaction initiated through the amine groups of analytes being tested. To ensure that the results observed are due to antibody-antigen interaction, rather than electronic drift or fluctuations, we deployed the third GFET as the comparative electronic control. The third graphene channel in this GFET was modified with Tw20 only, to serve as a blocking layer, with the expectation that it would not respond to introduction of any analyte into the solution. Each step of functionalization was characterized electrically (Figure2 c-d) and optically (Figure S3 and S4) with all devices assembled, showing a consistent trend indicating successful immobilization and blocking.

It is important to note that our chips are made for single use. To understand the effect of time on the stability of the chips we performed IV curve characterization using bare graphene GFET over a period (14 days) and recorded the movement of CNP (Figure S6). On an average the CNP value was found to be 135 mV with a standard deviation of 1.46 mV. In practical use, this drift can be accounted for in the electrical readout processing.

### Device testing and performance with pristine buffer (0.01X PBS)

To evaluate the sensing capability of the device, we performed a series of time trace measurements where all onboard transistors were exposed to varying concentrations of both COVID-19 S-protein (Spike) and Flu Hemagglutinin (HA) proteins at different intervals as outlined in the measurement protocol in (Figure S7).

The antigen-antibody interaction utilizes the uniform turbulent diffusion of viral proteins delivered^46, 47^ in low ionic strength PBS, entailing a facile operating procedure, where the user simply pipettes a drop of the viral protein solution onto the device and observes a response within seconds. Prior to testing the device against the target proteins, a negative control protein test was conducted with bovine serum albumin (BSA) as the analyte to verify its specificity (Figure S8). We established a precise dual detection of the two viral particles without cross-reactivity of the signals; hence each time the characterized devices were exposed to control proteins to study cross-reactivity and specificity.

For all the time-resolved trace measurements, the gate voltage was set to the value which exhibited the highest transconductance (V_gmax_) for the chip in PBS 0.01X post functionalization (Figure 3a). The gate voltage at the highest transconductance value generally ranged from 120mV to 200mV. This was carried out to ensure that the channel had the maximum sensitivity^46, 48^ to any activity on the surface of antibody-decorated graphene channels.

**Figure 3.**
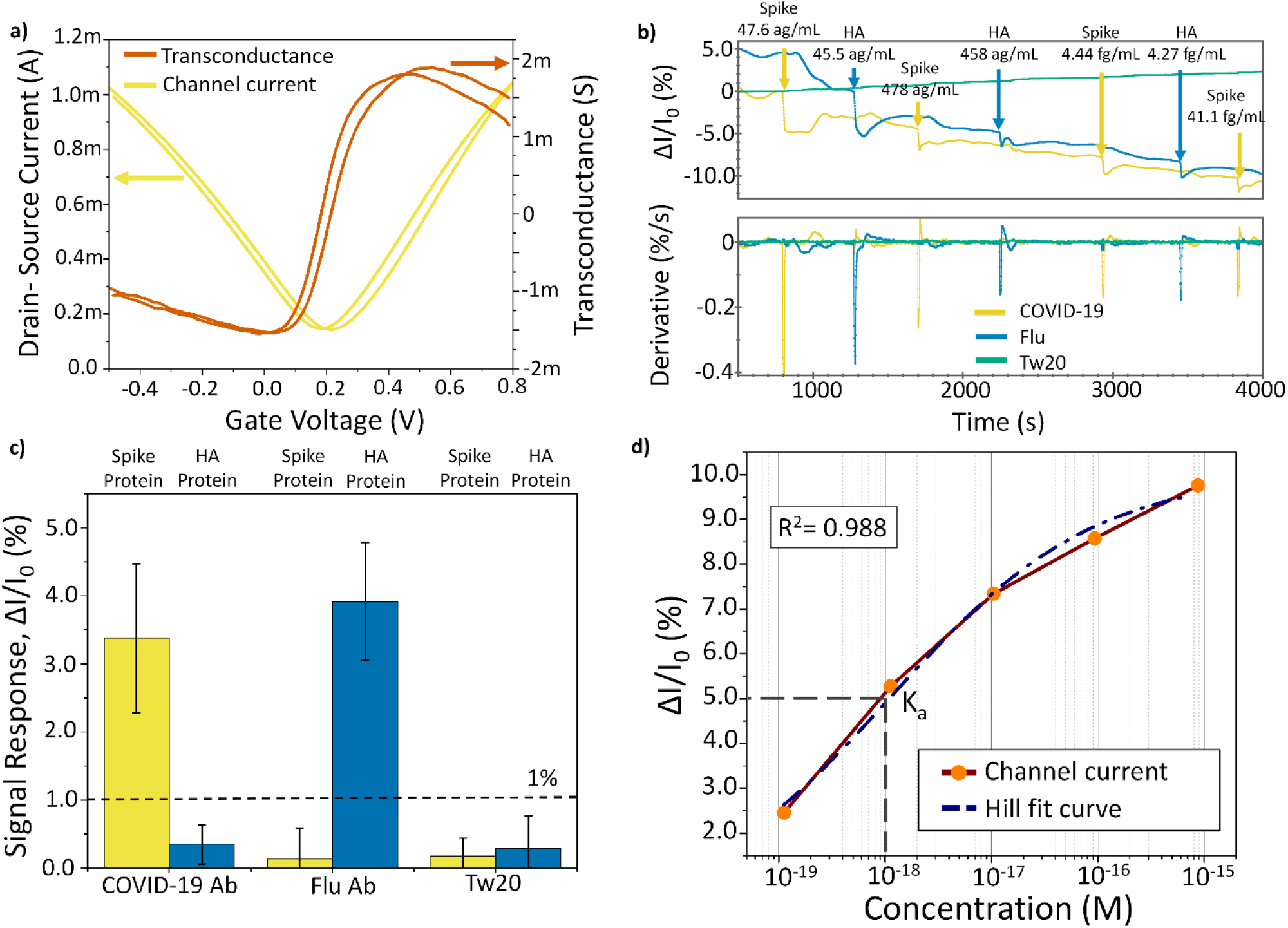
Simultaneous dual detection of COVID-19 (Spike) and Flu (HA) proteins. a) Transconductance curve (vermillion) of the antibody coated GFETs to verify the gate voltage at the point of highest transconductance to ensure the highest sensitivity. b) Time series measurement demonstrating simultaneous detection of both COVID-19 (yellow) and Flu (blue) along with the control (green) and their first derivatives on the same timeline indicating the exact moment of detection and differentiating from other event-induced artifacts. The antigens were introduced in successively increasing concentrations. c) Average signal response for the interaction with each antibody against Spike and HA across 4 devices at ∼50 ag/mL in PBS 0.01X buffer. A 1% threshold for signal response was assigned to differentiate a specific from a nonspecific antibody binding. d) Hill-fitted curve of the change in current of GFET immobilized with CR3022 antibodies vs. successively increasing concentration of Spike proteins. Association constant (K_a_ ∼ 1 × 10^-18^ M) extracted from the Hill-fit curve.

Figure 3b details the response of the quadruple architecture GFET chip to the introduction of both viral surface proteins. The proteins were serially diluted 10-fold with PBS 1X in maximum recovery microtubes (1.5 mL). Each dilution was then further diluted 100-fold into PBS 0.01X prior to being loaded onto the device. The first viral protein to be introduced was Spike protein with the lowest concentration (47.6 ag/mL), following which the channel current stabilized. After stabilization, the second viral surface protein, HA was added with the similar mass concentration as that of the first dosage of Spike protein. For each successive pair of additions, the concentrations of both the control proteins were kept similar. As expected, upon the introduction of Spike protein, the quarter functionalized with CR3022 registered an immediate change in conductance, leading to drop in the current while the GFET functionalized with FI6v3 experienced negligible change. Similarly, the introduction of HA induced a significant drop in channel current in the GFET functionalized with FI6v3 without inciting a significant reaction in the CR3022 GFET, underscoring the high specificity of the functionalization scheme. This can be further confirmed through the change in normalized channel current (ΔI/I_0_= (I_0_-I)/I_0_) observed for the first instance (first concentration at ∼ 50 ag/mL) of introduction for each protein, as shown in Figure 3c. The device design and measurement protocol produced reproducible results as the chip produced results with similar trend over 4 different devices. The mean change in normalized channel current as observed across the devices tested for COVID-19 GFET upon application of Spike protein is 3.37% (StD: 1%), while upon application of HA is 0.35% (StD: 0.28%). Similarly, upon introducing HA in the GFET with FI6v3, the change in normalized current is 4% (StD: 0.8%), while reaction of Spike protein had a minuscule change of 0.14% (StD: 0.45%). The significant difference in values indicates that the quadruple GFET architecture can successfully identify the control protein while preventing cross reactivity thus demonstrating the capability to function as both a sensitive and specific dual protein detector. The GFET passivated with Tw20 shows a minuscule change in the channel current, averaging at 0.3%, upon addition of any of the above-mentioned analytes, thus serving as a comparative electronic control, revealing the underlying variability in signal without interaction with the biological media. Choosing a cutoff of 1% for the first concentration of the antigens tested we capture 100% of true positives and reject 100% of cross GFET and chemically passivated GFET. 1% change in the normalized signal was chosen as the thresholding value to declare a true positive amongst all the 4 devices since it encompassed the maximum change in normalized current value for cross reactivity observed amongst all the devices (0.9%) while also being 4 times the mean (Figure 3c) response deduced for cross GFET reaction for the first tested concentration.

The derivative of the time series curve eliminates the impact of drift and other electronic artifacts observed in the real-time traces, as shown in Figure 3b, serving to accurately distinguish the instances of introduction of either Spike or HA protein from other artifacts in the measurements. As observed in Figure 3b, response to the first dosage of Spike and HA recorded the most significant drop in source-drain channel current in their respective GFETs in comparison with the successive drops in current observed at later dosages. The amplitude of the change in channel current decreases with an increase in the dosage of the protein. To understand the trend observed in channel current upon addition of successive higher concentrations of protein, kinetics of the antigen-antibody at the graphene interface was examined. The dissociation constant (K_d_) is extracted from the ΔI/I_o_ vs. Spike protein concentration Hill-Langmuir model (Eq. 1) fitted protein concentration curve as shown in Figure 3d,

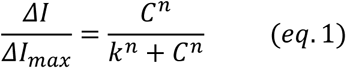

The K_d_ value obtained through the Hill fitted (Eq. 1) data points is 0.147 nM, and the Hill coefficient (n) stands at 0.45. The Hill coefficient below 1 indicates that the interaction between the antigens and the antibodies follows negative cooperative binding^49^. This implies that the first instance of interaction between the antigen and the antibodies is the strongest, while the reaction at successively increasing concentrations is likely blocked by the presence of viral surface proteins already interacting with antibodies near the surface, leading to a diminished signal response.

When analyzing the device performance, we observed overall sensitivity of the devices is very high, above other emergent technologies. Sensitivity was calculated by performing a linear fit on the linear range of the (I/I_0_) % vs log(M) curve, achieving 2.4% change in signal per log(molar) concentration for COVID-19 (2.4%/log (M)) and 1.9% change in signal per log(molar) concentration (1.9%/log(M)) of Flu (Figure 4a). Such sensitivity levels provide superior resolution for detecting and quantifying analytes at extremely low concentrations. Although we report our experimental LoD, practically, the low noise level of our system suggests we could detect down to concentrations of tens of viral surface proteins per mL via single-molecule interactions with the surface^57^. Apart from the high sensitivity, the devices come with a rapid response time of around ∼10s after addition of analyte (Figure 4b), which is amongst the fastest response times reported by any platform^7^. Finally, effective binding between the antibodies and antigens can indeed occur rapidly ^58^ (Supplementary note 2), as it has been corroborated by multiple methods, including Bio-Layer interferometry^59, 60^. There is also a recent report of a GFET-based devices which show a response time within the same time range^23^.

**Figure 4:**
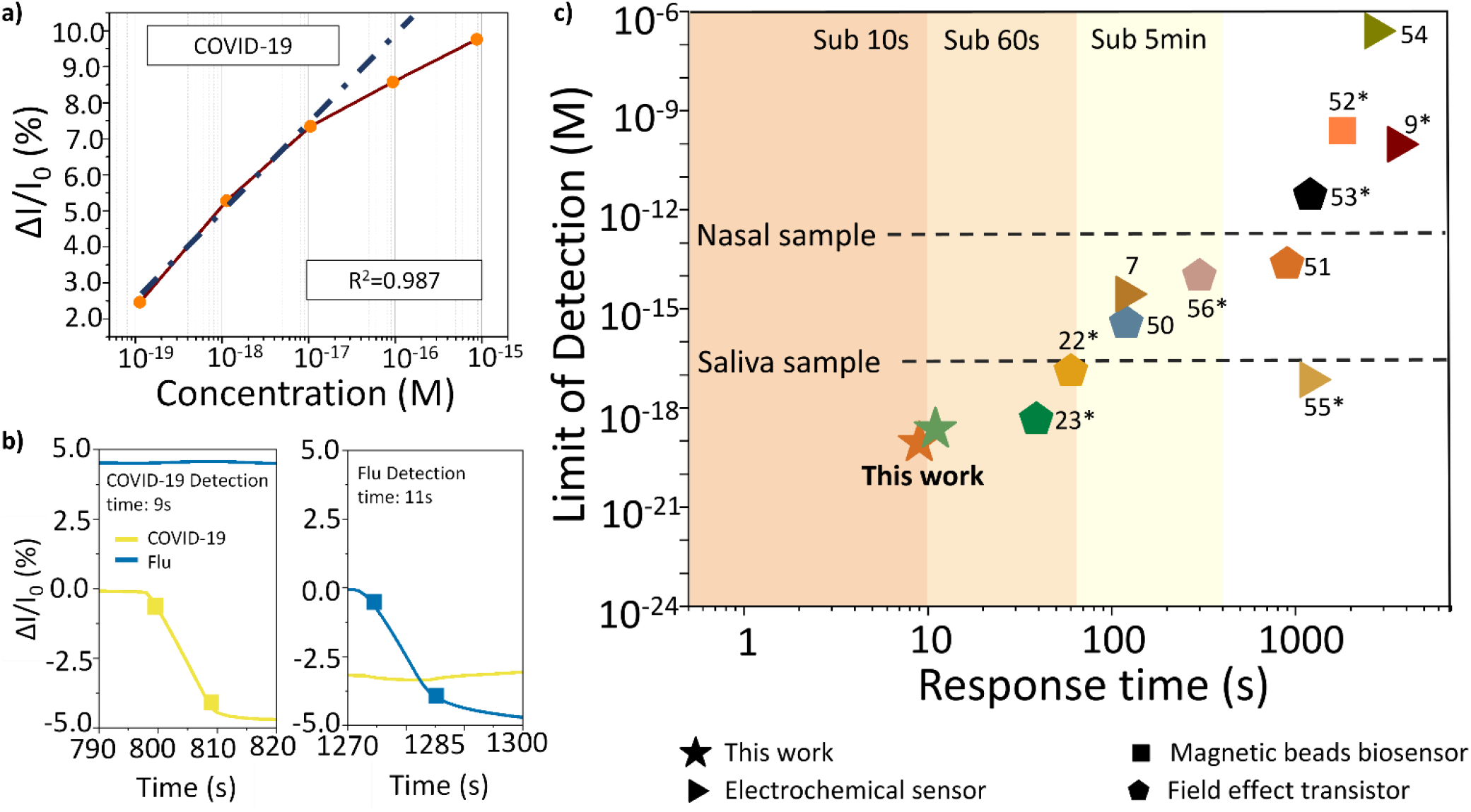
Device sensitivity, time response, and device performance in artificial saliva. (a) Sensitivity of the GFET for COVID-19 through fitting (blue dotted fitted curve) of the linear part of the concentration vs. current response curve (brown curve with orange data points). (b) Time response of the device upon introduction of both COVID-19 and Flu proteins. (Yellow curve is for COVID-19 and blue curve for Flu). The squares on each curve mark the 10% and the 90% of the step response that occurs due to change in channel current upon interaction of the specific antigen with the antibodies. (c) Comparative LoD with contemporary technologies. Benchmarking chart with LoD and response time of the current state-of-the-art technologies available for SARS-CoV-2 diagnoses. The dashed lines represent the minimum LODs required for different types of samples for successful detection. (This work: green star is for HA, and the red star is for Spike protein.) * Recalculated molarity using the molecular weight provided in the referred work. ^7, 9, 22, 23, 50–56^

This instantaneous turnaround time, if productized, could be particularly useful in locations with high patient load. Based on the experimental data, the device response’s standard deviation (σ) level is 0.04% and 0.07% for COVID-19 and Flu, respectively. Since our lowest detected concentration response is well above the 3σ or even 9σ value for both COVID-19 and Flu at an average of 3.37% and 4%, respectively, we have an experimental LoD (88 zM) 5-fold lower than the previously reported LoD (Figure 4c) for detection of COVID-19^23^. Amongst other technologies like electrochemical sensors^55^, reporting similar LoDs (Table 1), our device demonstrates the fastest turnaround time while also presenting an inexpensive electronic alternative. Moreover, we performed an additional experiment demonstrating the device’s capability in operating with clinically relevant saliva-based sample (Supplementary note 1). The results obtained from the experiment, expectedly, follow the same pattern, validating applicability of the reported cross-functionalization (Figure S9). For this preliminary data the threshold for signal response chosen for assigning specific and non-specific reaction was 0.6%. This threshold is 4 times more than the highest change in signal observed for non-specific reaction between CR3022 and HA. The preliminary response observed through the experiment with complex medium along with experimental data obtained with PBS 0.01X medium across 4 devices strongly points towards the device’s capability of performing reliably with patient samples as well. Evaluating clinical samples as demonstrated by Seo et al.^22^ with comprehensive statistical analysis is a matter of future studies.

**Table 1.** Summary of the existing antigen/antibody testing platforms. (^Š^ – recalculated to molarity using molecular weight provided in the referred paper).

| Method | Response time | LoD |  | Multi-variant | Diagnostic Biomarker | Ref. |
| --- | --- | --- | --- | --- | --- | --- |
|  |  | Copies or g/mL | M |  |  |  |
| **Graphene FET** | ~10s | ~50 ag/mL | 88 zM (S protein)<br>227.2 zM (HA) | **Yes** | COVID-19 and flu viral proteins | **This work** |
| FET | 2 min | N/A | 0.37 fM | No | COVID-19 viral RNAs | 50 |
|  | 1 min | 30 copy/mL | N/A | No | COVID-19 viral DNA and RNA | 24 |
|  | 38.9 sec | 35 ag/mL | 0.458 aM <sup>§</sup> | No | COVID-19 viral proteins | 23 |
|  | 20 sec | 25 pg/mL | N/A | No | COVID-19 viral protein | 10 |
|  | 1 min | 1fg/mL | 13.07 aM <sup>§</sup> | No | COVID-19 viral protein | 22 |
|  | 5 min | 0.55 fg/mL | 7.18aM <sup>§</sup> | No | COVID-19 viral protein | 56 |
| OECT | 10min | N/A | 23 fM | No | COVID-19 viral protein | 51 |
|  | 20 min | 0.36 fg/mL | 2.78 pM <sup>§</sup> | No | COVID-19 viral proteins and virus specific antibodies | 53 |
| LAMP | 35 min | 1000 copies/mL | N/A | No | COVID-19 viral RNA | 64 |
|  | 20 min | 10000 copies/mL | N/A | No | COVID-19 viral RNA | 65 |
| Electrochemical | 6 min | 229 fg/mL | N/A | No | COVID-19 viral protein | 66 |
|  | 60 min | N/A | 1 fM | No | COVID-19 virus | 67 |
|  | 60 min | 2.9 ng/mL | 96.6 pM <sup>§</sup> | No | COVID-19 viral protein | 9 |
|  | Real Time | 200 copies/mL | N/A | No | COVID-19 RNA Genome | 68 |
|  | 2 min | N/A | 2.8 fM Spike<br>16.9 fM Abs | No | COVID-19 Antibody | 7 |
|  | 10 min | 0.1 µg/mL | N/A | Potentially yes | COVID-19 viral protein and virus specific antibody | 69 |
|  | 45 min | N/A | 260 nM | No | COVID-19 viral protein | 54 |
|  | 30 min | 19ng/mL | 24.7 nM <sup>§</sup> | No | COVID-19 viral protein | 55 |
| Paper based sensor | 45 min | 1 µg/ml | N/A | No | COVID-19 antibody | 70 |
| Surface Plasmon Resonance | 30 min | N/A | 2 nM | Yes | RNA genome | 71 |
|  | 10 min | 0.18 µg/mL | N/A | No | RNA genome | 72 |
| Phase Molecular Recognition | 5 min | N/A | 48 fM | No | COVID-19 viral protein | 73 |
| Magneto sensor | 30 min | 8 ng/mL (S)<br>19 ng/mL (N) | 0.24 nM<sup>§</sup> | No | COVID-19 viral proteins | 52 |

Our device’s high sensitivity and low experimental LoD can be attributed to the deployment of low strength ionic buffer and PEG-NH_2_ in functionalization to combat the screening effect caused by short Debye length in high ionic strength buffers. Aiding the specific functionalization scheme is also the selection of the most sensitive V_gs_ corresponding to a high transconductance value. By virtue of the linear relationship (Eq. 2) between transconductance and W/L ratio, the high W/L ratio of 7.5 in the device architecture enables higher transconductance, imparting higher sensitivity in turn translating to ultra-low LoD.

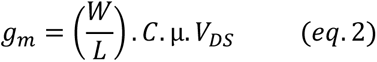

Our device standing at 88 zM is already approaching breath sample detection levels (118.2 zM)^61^ (Supplementary note 3) while already surpassing the minimum LoD requirements for nasal (163 fM) and saliva sample (16.3 aM)^62, 63^. Such low LoD, as exhibited by our device, allows versatility in selecting the type of sample and can potentially reduce the time for administering the test after exposure.

Owing to their molecular weights, theoretically, the lowest possible concentration with Spike and HA protein is ∼1.67 zM. Our device’s lowest measured concentrations indicate the capability of almost approaching single molecule detection for each viral protein in their respective GFETs with essentially an immediate turnaround time.

### Conclusion

Designing for simultaneous and differential detection of COVID-19 and Flu, we describe a sensor platform consisting of an array of GFETs driven through a common gate and shared biological media with LoD at 88 zM for COVID-19 and 227 zM for Flu. The outstandingly low detection limit is attributed to be due, in part, to photolithography contamination-free graphene device quality since the fabrication protocol does not include any on-wafer photoresist or plasma etching steps. Such clean surfaces are expected to afford more uniform antibody coverage and better contact between the active region (graphene) and the electrode. In addition, our devices are passivated with PEG, which is known^44, 45^ to uplift the Debye screening layer from graphene surface, and by using low-molarity buffer solutions, we ensure that the proteins attach rapidly and with high sensitivity. These findings provide a proof-of-concept solution to the problem of rapidly differentiating two or more diseases with overlapping symptoms. The device enables immediate readout with a rapid response time of around 10s which captures the quick binding that antigens and antibodies undergo under ideal conditions (Supplementary note 2). The differential sensing results from high specificity and sensitivity accorded by the specific immobilization of the antibodies on two GFETs accompanied by an electronic control in the form of passivated GFET. Unlike the paper based PoC solutions, our platform presents a highly specific, facile, and portable electronic point of care technology which would critically benefit in the areas with high density and volume of patients and visitors such as clinics, nursing homes, universities, mitigating the bottlenecks created due to high turnaround times and complicated testing procedures. The multi-channel GFET device is also highly versatile since it can be repurposed with antibodies/receptors specific to other diseases, thus serving to track and mitigate future epidemic and pandemic threats.

## Methods

### ELISA Protocol

High binding 96 well plates [Costar cat 07-200-721] were coated at 2 ug/mL with S protein or HA overnight at 4 °C. Plates were washed three times with PBS 1X with 0.05% TW-20 (PBST) and were blocked with PBS 1X, 2% skim milk for 2 hours at room temperature. Antibodies in (1X or 0.01X) PBS, 0.05% TW-20, and 1% skim milk (PBSMT) were serially diluted across the 96 well plate before a 1-hour incubation. Goat Anti-Human-IgG with horseradish peroxidase (HRP) (Sigma-Aldrich^TM^ cat A0293) diluted 1:5000 in PBSMT 1X was used as a secondary antibody and incubated for 30 minutes. 1-Step^TM^ Ultra TMB-ELISA Substrate (Thermo Scientific^TM^ cat 34029) was used to develop the plates and the reaction was quenched with 2M H2S04. Absorbance values were measured at 450 nM on a Synergy H1 microplate reader (BioTek^TM^).

### Proteins

Gblocks ordered from Integrated DNA Technologies (IDT) containing antibody variable heavy or light chains were inserted into mammalian expression vector pcDNA3.4 by Golden Gate cloning and validated with sanger sequencing. Antibodies were expressed using the Expi-293^TM^ Expression System (Thermo Scientific^TM^ cat A14635) and purified with Pierce^TM^ Protein G Plus Agarose (Thermo Scientific^TM^ cat 22851). A stabilized version of the S protein, Hexapro was expressed using the Expi-293 expression system and purified using Ni-NTA agarose (Qiagen cat 30210). All proteins produced in house were validated on SDS-PAGE gels and quantified using the Pierce^TM^ Coomassie Plus (Bradford) Assay Kit (Thermo Scientific^TM^ cat 23236). Proteins purchased commercially included the HA strain H3N2 A/Singapore/INFIMH-16-0019/2016 (Native Antigen) and powdered BSA (Thermo Scientific cat BP9706100).

### Device preparation

Shadow mask was employed to deposit gold on Si/SiO_2_ wafer as three terminals to create a 4-GFET array structure of the device. Cr/Au (10nm/90nm) layers were deposited through e-beam deposition and lift off techniques. Wet transfer method was utilized to transfer graphene onto the substrate.

Commercially obtained graphene sheet grown on copper (Grolltex) was spin-coated with Poly (methyl methacrylate) (PMMA) (PMMA 950 A4, MicroChem). After spin coating, the PMMA/graphene/Copper stack was baked at 150 ℃ for 10 minutes. The PMMA/graphene/Copper stack was upturned with the Cu side exposed and was subjected to Oxygen plasma for 30 sec at 30% flow rate. The copper sheet with PMMA/graphene film was then cut into 10mm x 10mm pieces and placed into Ammonium Persulphate, (NH_4_)_2_S_2_O_8_, for 24 hours to dissolve the copper. Pieces were placed with PMMA side facing upwards to allow the copper to dissolve. PMMA/graphene film pieces were rinsed and allowed to soak in deionized (DI) water for a total of three consecutive times and then transferred to the silicon wafer with a gold deposit. PMMA/graphene transferred wafers were left to air dry overnight and then baked at 150 ℃ for 10 minutes. Wafers were then placed in an acetone bath for 24 hours to dissolve the PMMA layer. Bare graphene wafers were rinsed in ethanol and DI water and then dried with the air gun. Dried wafers were baked at 150℃ for 10 minutes. PDMS enclosures were made by cutting rectangular pieces of PDMS and using liquid PDMS to hold them together. The outer PDMS boundary was made with a taller height than the inside cross enclosure to allow overflow between channels on the top (during measurements) of the inside but to prevent leakage to the outside. Inter-leaking between channels was tested using isopropyl alcohol. Small lengths of copper wires were stripped at both ends and connected to the common source, the drain, and the ground through contact with the gold layer on the device and the use of silver epoxy (MG Chemicals 8331S Silver Epoxy Adhesive) to make sure the wires stayed attached to the device.

### Functionalization

10mM PBASE (Anaspec, AS-81238) solution in Dimethylformamide (DMF) (Thermo Scientific, 20673) was prepared. PBASE and DMF solution was added to both the COVID-19 and Flu-designated GFETs. Glass slide cleaned with ethanol was placed over the device during the 1-hour incubation period to mitigate the risk of DMF evaporating. Starting with one GFET at a time, the PBASE/DMF solution was taken out, and the GFET was rinsed with plain DMF once and DI water three times. Rinsing was performed quickly to avoid drying out the GFET. 50 ug/mL of COVID-19 (CR3022) antibodies were added to the GFET and incubated for an hour. Simultaneously, the Flu-designated GFET went through the same rinsing steps with DMF and DI water with 50ug/mL of the Flu antibodies, FI6v3, being added with the same incubation time. After one hour of incubation, CR3022 and FI6v3 were taken out of GFET one at a time, and GFET was rinsed with PBS 1X three times. After the rinse, 3mM PEG-NH_2_ (Broadpharm, P-22355) and PBS solution were added to the GFET and incubated for another hour. 1M ETA (Sigma Aldrich, 110167) solution was prepared by combining ETA with PBS 1X (pH8). After both GFETs had been incubated with PEG-NH_2_ for an hour, PEG-NH_2_ inside the GFET (one GFET at a time) was dispensed and rinsed with PBS 1X three times. The prepared solution of ETA was placed into the GFET and incubated for another hour. All ETA steps were repeated for the other GFET with antibodies. Tw20 (Sigma Aldrich) was placed into a third GFET that didn’t contain any antibodies as a negative electronic control. After an hour of incubation with ETA, the ETA solution was dispensed from the GFETs with antibodies and rinsed with PBS 1X. Tw20 was also taken out of its designated GFET, and the GFET was rinsed with PBS 1X.

### Characterization

To ascertain the presence of PBASE and other functionalization reagents on graphene, Raman spectroscopy was performed using Witec Micro-Raman Spectrometer Alpha 300. Electrical functionalization was carried out using Keithley B2902A.

### Dilution of Protein samples for measurements

Proteins from a frozen stock previously quantitated by Bradford assays and Nanodrop Spectroscopy were serially diluted 10-fold with PBS 1X in maximum recovery 1.5 mL microtubes (Axygen MCT-150-L-C). Each dilution was further diluted 100-fold into PBS 0.01X prior to being loaded onto the device.

### Device measurements

Device measurements were carried out using Keysight B2909 A source-meter for both I-V curve and time-resolved measurements. For functionalization step I-V curves, the PDMS chamber was filled with PBS 1X, and the gate voltage was swept over a range of -0.3 to 0.7 V with V_ds_ = 0.1V. For time series measurements against the proteins, the PDMS chamber was initially filled with PBS 0.01X at 400ul and activated with the chosen gate voltage (voltage for highest transconductance) and V_ds_= 0.1V. The chip was allowed to stabilize for at-least 500s. Before introducing the proteins of interest, a third-party test with Bovine Serum Albumin (BSA) was conducted by adding 25ul of the BSA solution into the PDMS well. After the test, the chip was disconnected from the source meter and thoroughly rinsed and refilled with PBS 0.01X and reconnected to the source meter with the V_gs_ and V_ds_ set at the same value as previously stated. Once the reconnected chip stabilized, protein samples were introduced at different concentrations. The samples of both Spike and HA proteins were prepared through serial dilution in PBS 1X. Since the buffer being used for testing is PBS 0.01X, the stock proteins prepared in PBS 1X were resuspended in PBS 0.01X (adding 10ul of protein in 1X PBS into 990ul of 0.01X PBS) and thoroughly mixed 5 seconds prior to introducing them to the chip (25uL of the protein in 0.01X PBS added to 400uL PBS 0.01X solution on the chip). The measurement was performed in pairs, the first 25ul of Spike protein in PBS 0.01X was introduced into the chip. Once the current stabilized after reaction in the COVID-19 GFET, then 25ul of HA protein in PBS 0.01X was added to the chip. This procedure was performed for each concentration of protein to be tested.

## Corresponding Author

*

## Author Contributions

DK, DA, and AE devised the project and experimental plan. DK, NK, CG, and SM performed device fabrication. NK, DK, CG, and SM assembled devices, performed chemical functionalization, measurements, optical and electrical characterization, and data analysis. DT prepared biological samples for functionalization and testing. DT performed biochemical assays to characterize reagents. NK, DK, DT, CG, and DA interpreted results. NK, DK, DT, CG, and DA wrote and edited the manuscript.

## Notes

The authors declare no competing interests.

## Data Availability

All data produced in the present work are contained in the manuscript

## Acknowledgements

This work was supported in part by the National Science Foundation, Awards #2033846 and #2222907, and the University of Texas ECE departmental research funds. D.A acknowledges the Temple Foundation Endowed Professorship and the Jack Kilby Endowment. We also acknowledge the efforts by Jo Wozniak for the Table of content figure. We also acknowledge GrollTex for their high-quality Graphene sheets. N.K. acknowledges her colleagues Sanjay Rajendran and Sudhanva Vasistha for their inputs for the graphs and figures. D.A acknowledges Dr. Oluwadamilola Oshin of Covenant University, Nigeria, for initial work on functionalized graphene transistor biosensors at UT-Austin. The authors appreciate the sensor and measurement insights provided by Brett R. Goldsmith and Francie Barron of Cardea Bio.

## Notes

### Competing Interest Statement

The authors have declared no competing interest.

### Summary of Updates

Affiliation updated; figure 1 revised; main manuscript modified; supplementary updated

