## Supplementary Information for "Graphene Field Effect Biosensor for Concurrent and Specific Detection of SARS-CoV-2 and Influenza"

for

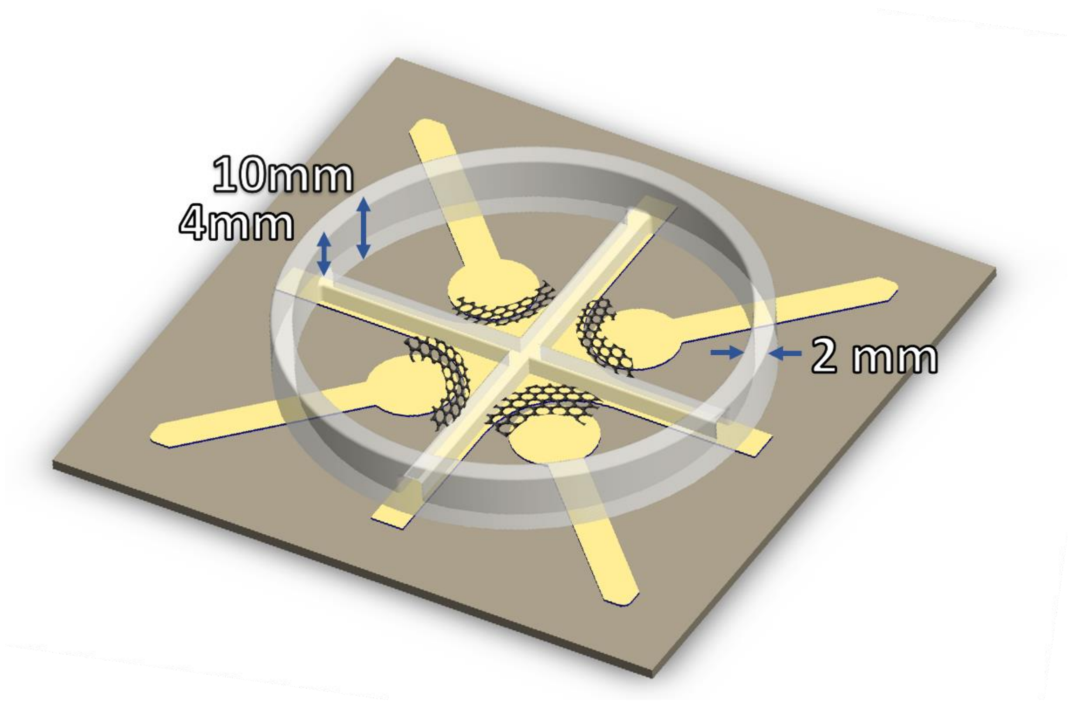


*Figure S1. Height difference between the inside PDMS cross enclosure and the outer enclosure. The height difference of the outer and the inner enclosure allows independent functionalization, while during testing it holds a common medium across all the GFETs, thus doing away with the requirement of complex microfluidics to enable concurrent detection.*

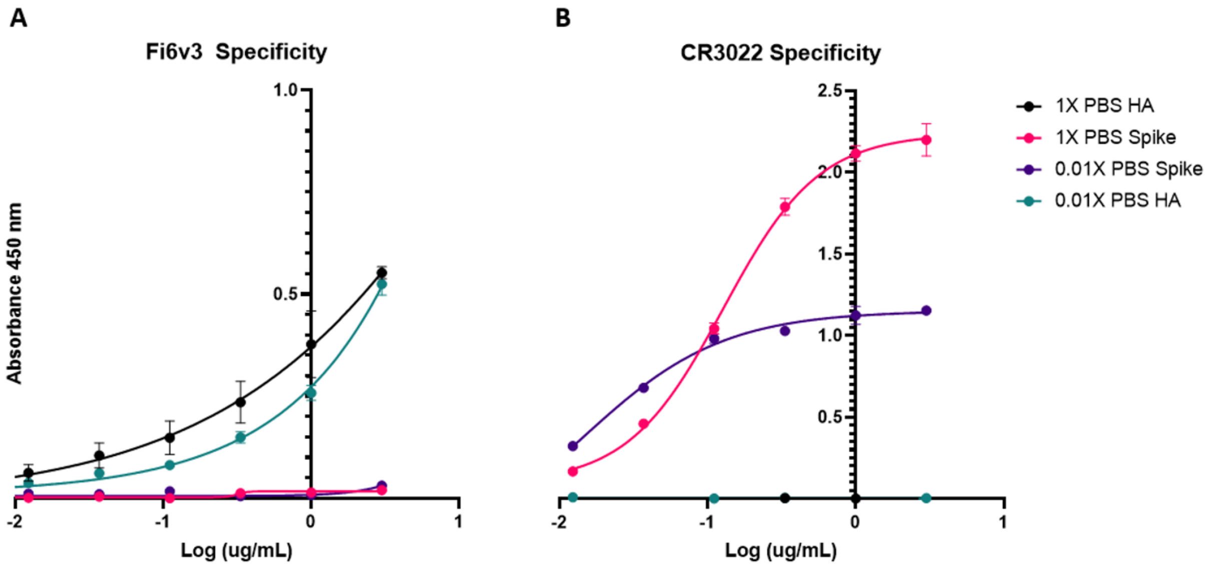


*Figure S2. Comparison of performance and specificity of the COVID-19 and flu antibodies in buffers of different ionic strengths a) Fi6V3 and b) CR3022 at 1X and 0.01X PBS.*

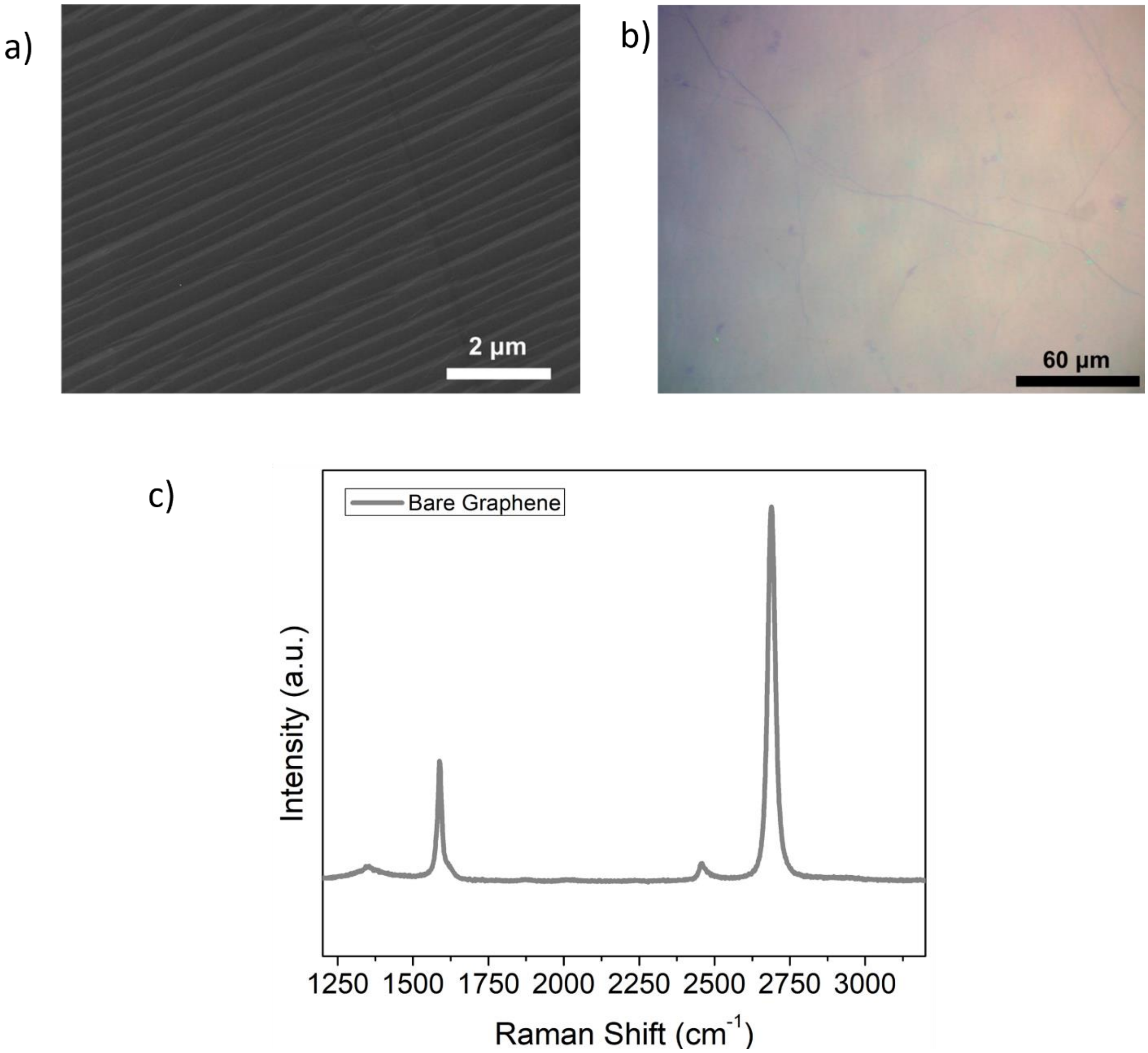


*Figure S3. Material characterization data of commercially obtained Graphene (from GrollTex) used for the experiments. (a) SEM image. (b) Optical image and (c) Raman data of bare graphene.*

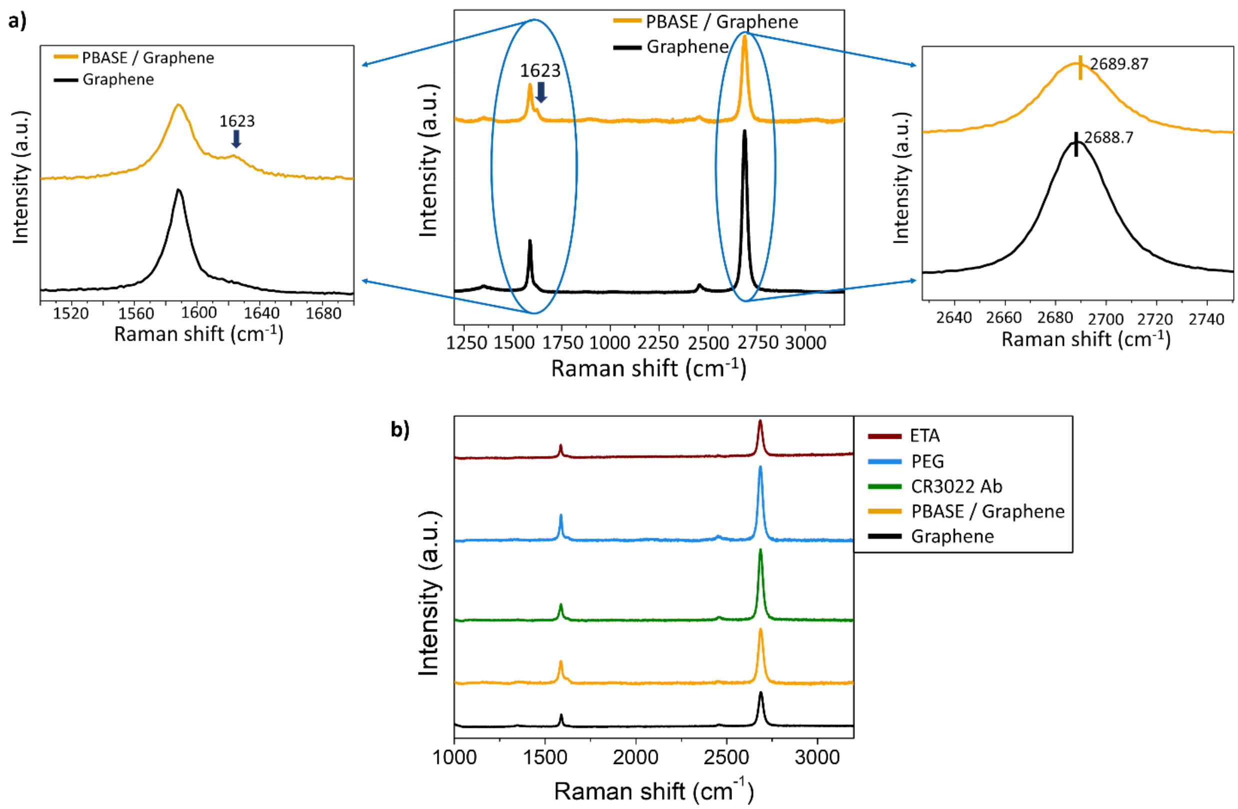


*Figure S4: Raman results of the all the steps of functionalization. (a) Full spectrum Raman spectroscopy after modification of bare graphene with PBASE. The presence of D' peak at 1623 $cm^{-1}$ indicates presence of pyrene resonance. The movement of the 2D peak to right by 1.3 $cm^{-1}$ indicates hole doping (b) Raman spectroscopy results of all the steps of functionalization.*

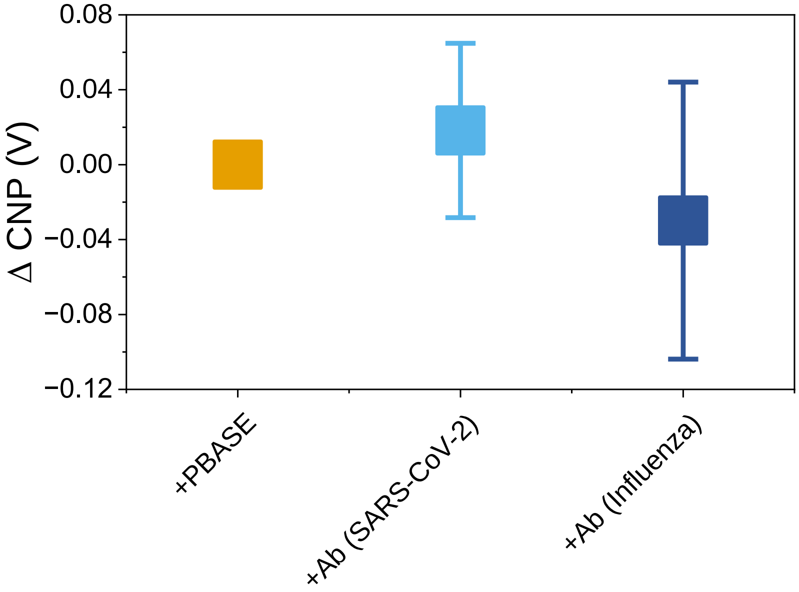


***Figure S5.*** *Change in Charge Neutrality Point (CNP) with respect to PBASE stage due to doping induced by the SARS-CoV-2 and Influenza antibody immobilization. Whiskers are ±SD.*

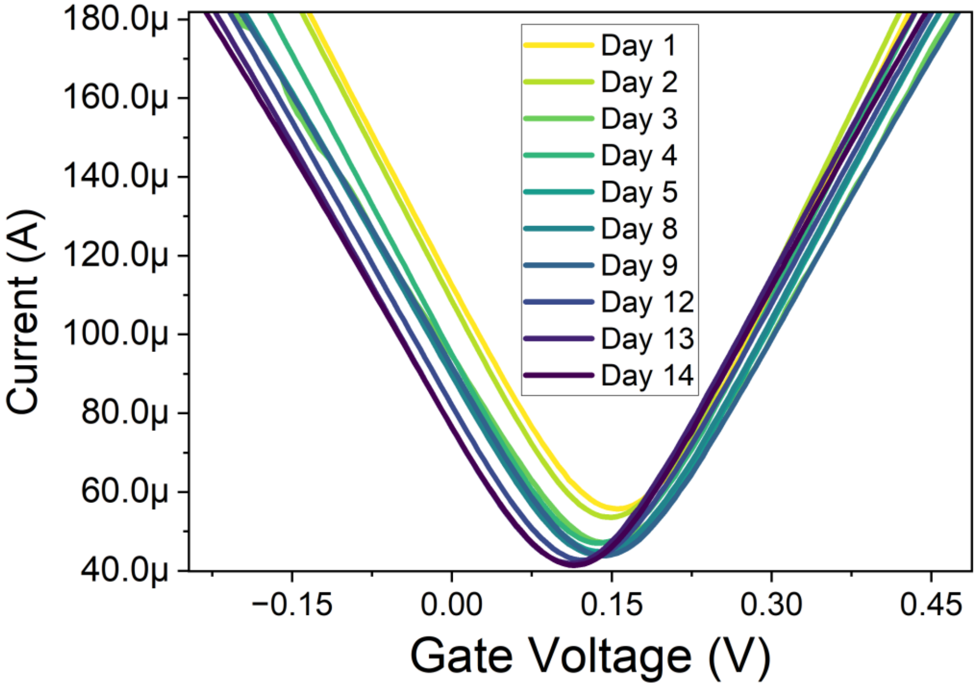


***Figure S6.*** Stability: Change in the CNP value of the bare FET over 14 days. Movement of the CNP value over a period of time (14 days).

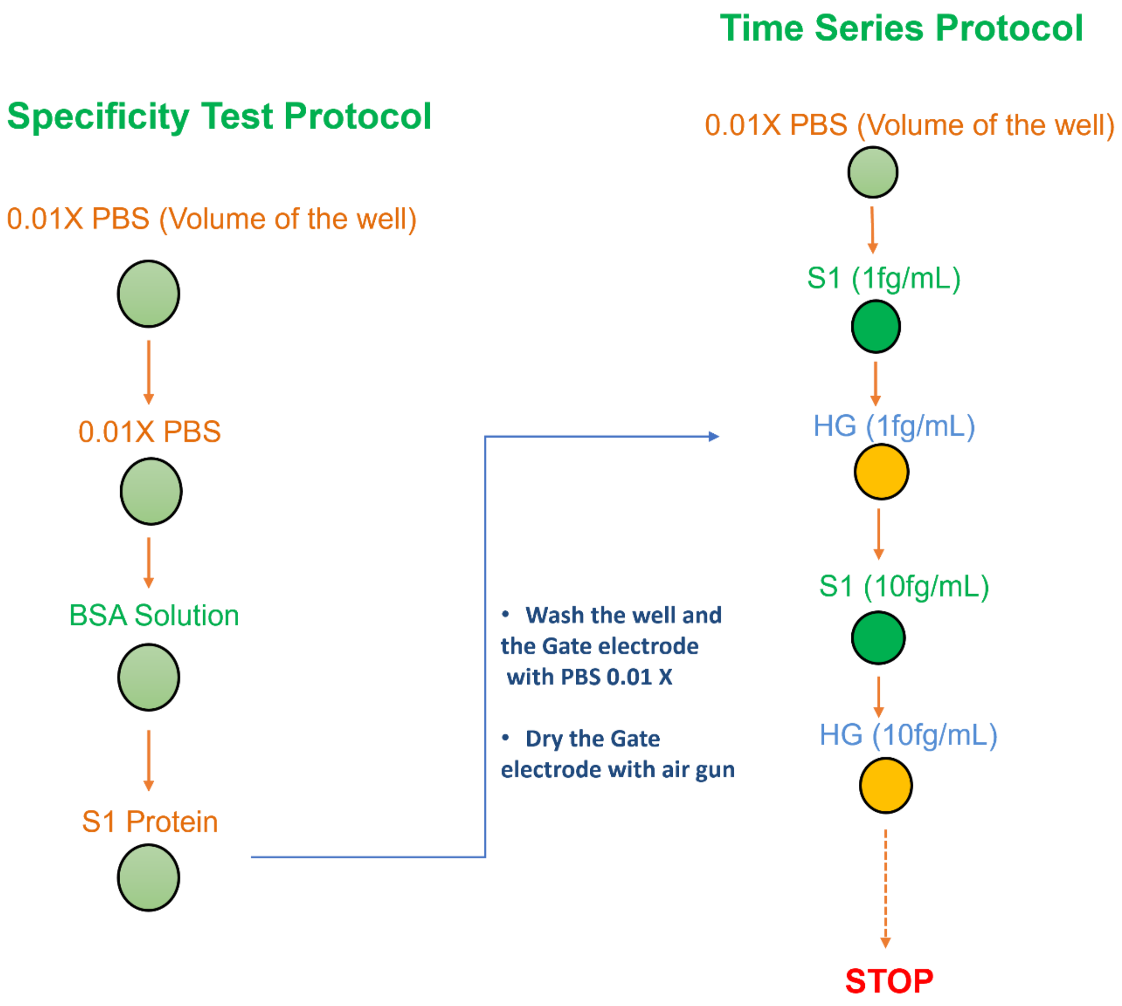


*Figure S7. Time series measurement protocol employed while testing the device against both COVID-19 and Flu antigens.*

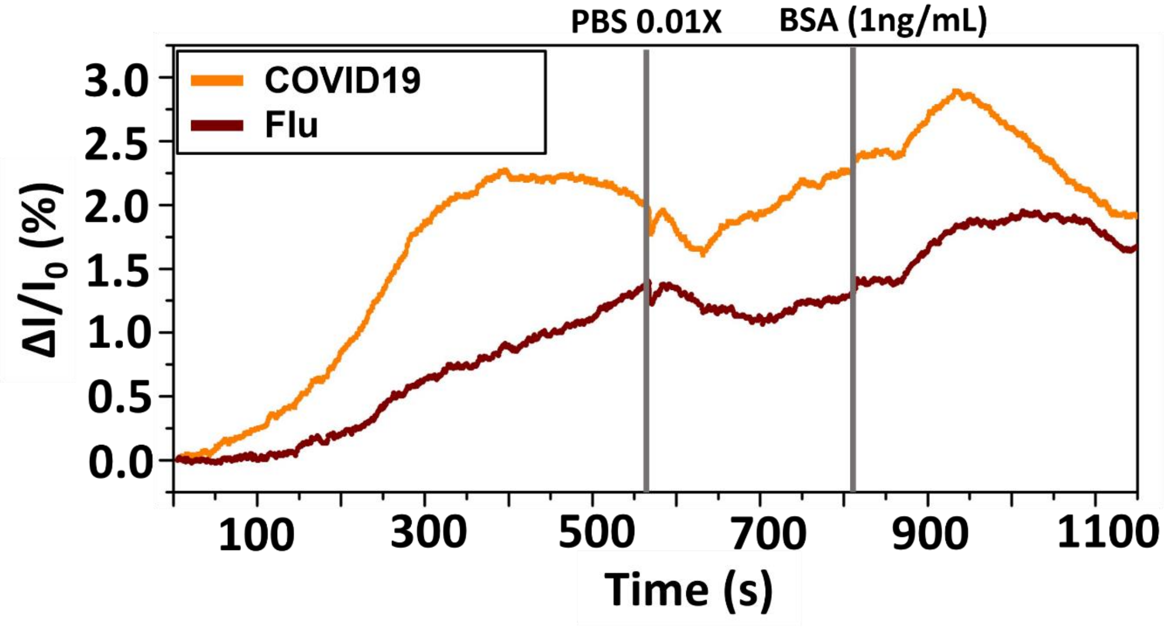


*Figure S8. Time series specificity test of the device against blank sample and negative control i.e., BSA.*

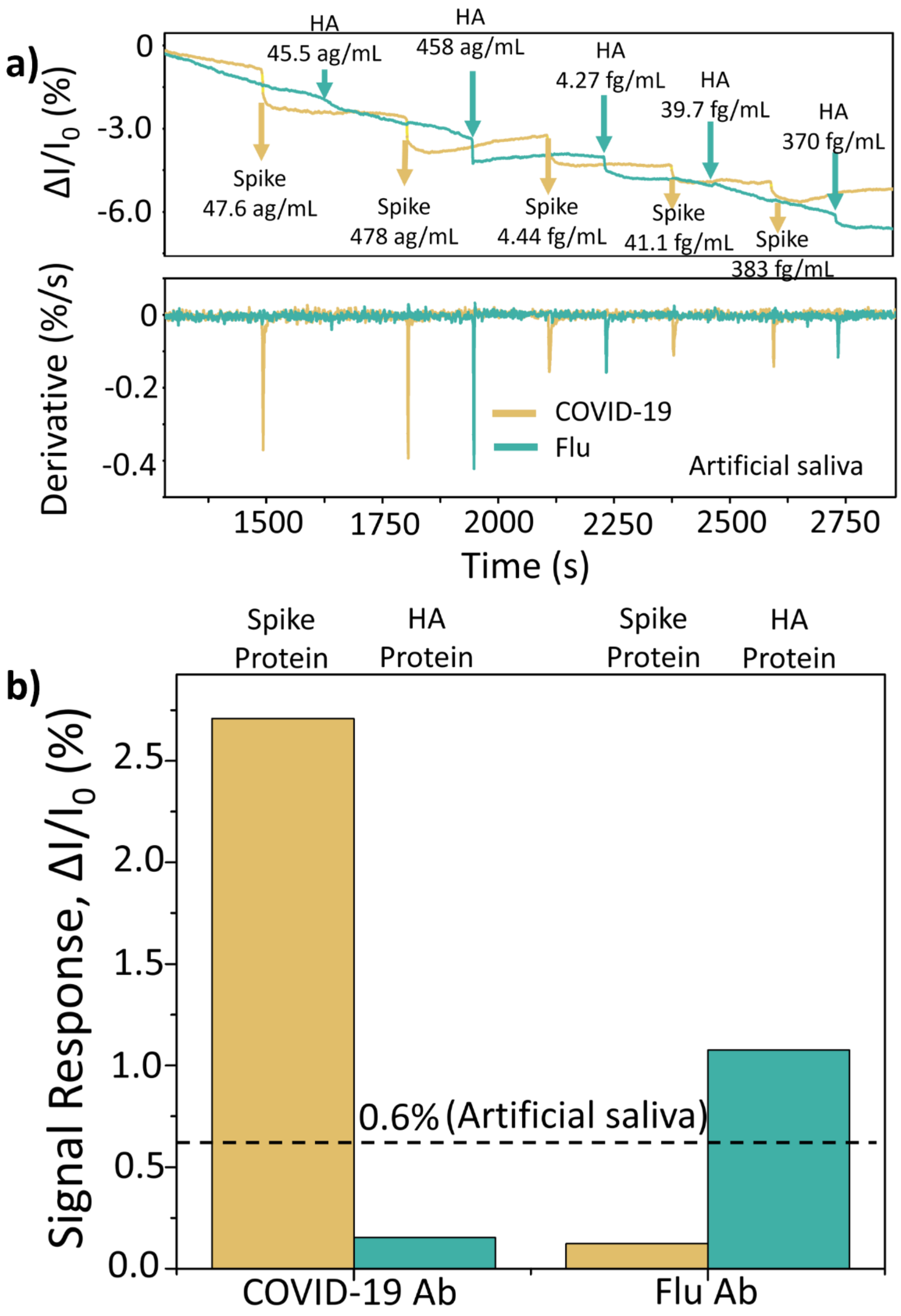


***Figure S9:*** *Device performance in artificial saliva. (a) Time series measurement demonstrating simultaneous detection of both COVID-19 (yellow) and Flu (blue) in artificial saliva and their first derivatives on the same timeline indicating the exact moment of detection and differentiating from other event-induced artifacts. The antigens were introduced in successively increasing concentrations. (b) Signal response for the interaction with each antibody against Spike and HA across devices at ~478 ag/mL. A 0.6% threshold for signal response was assigned to differentiate a specific from a nonspecific antibody binding, which is 4 times more than the* response *deduced for cross GFET reaction for the second tested concentration.*

**Supplementary note 1**

**Application in Complex Samples**

To demonstrate clinical applicability the device performance in a complex biological medium, it was tested against surface protein samples prepared in artificial saliva (AS). Following the similar operation protocol as followed in our initial measurements in PBS buffer, Figure S9a details the response of the quadruple architecture GFET chip to the introduction of both viral surface proteins. The proteins were serially diluted 10-fold with AS 1X in maximum recovery microtubes (1.5 mL). Each dilution was then further diluted 100-fold into PBS 0.01X prior to being loaded onto the device. Following the same measurement protocol, we first introduced the Spike protein with the lowest concentration (47.6 ag/mL), following which the channel current stabilized. After stabilization, the second viral surface protein, HA was added with the similar mass concentration as that of the first dosage of Spike protein. For each successive pair of additions, the concentrations of both the control proteins were kept similar. As expected, upon the introduction of Spike protein, the quarter functionalized with CR3022 registered an immediate change in conductance, leading to drop in the current while the GFET functionalized with FI6v3 experienced negligible change. Similarly, the introduction of HA induced a significant drop in channel current in the GFET functionalized with FI6v3 in most of the instances without inciting a significant reaction in the CR3022 GFET, holding high specificity even in a complex medium like artificial saliva. For this experiment the second dosage of ~ 478 ag/mL was chosen as the benchmarking concentration to observe the cross reactivity because the change in current upon introduction of the first HA concentration did not get registered in the Flu GFET. The change in normalized channel current as observed across the devices tested for COVID-19 GFET upon application of Spike protein is 2.27%, while upon application of HA is 0.15% (Figure S9b). Similarly, upon introducing HA in the GFET with FI6v3, the change in normalized current is 1.07%, while reaction of Spike protein had a minuscule change of 0.12%. The measurement data in artificial saliva shows significant difference in cross reactivity values thus establishing the operational capability of the device even in complex samples. A cutoff of 0.6% (4 times higher than the highest change in normalized current observed for non-specific reaction) was chosen for the second concentration of the antigens tested to capture 100% of true positives and reject 100% of cross GFET. Like the results observed in PBS buffer data, derivative of the time series curve was performed to eliminate the impact of drift and other electronic artifacts observed in the real-time traces, as shown in Figure S9a, to accurately distinguish the instances of introduction of either Spike or HA protein from other artifacts in the measurements.

**Supplementary note 2**

The "immune reaction" for the formation of an antibody-antigen complex involves non-covalent interactions such as hydrogen bonding and electrostatic charge pairs that can allow for the rapid formation of an antibody-antigen complex and a slower reversible dissociation. For the common characterization of kinetics for antibody-antigen complex formation, technologies such as Bio-Layer Interferometry (BLI) or Surface Plasmon Resonance (SPR) are regularly used to determine association and dissociation constants for the formation of a complex. Like our device, these technologies first coat antibodies to a surface that will then be monitored for changes in a selected property such as refractive index for SPR or wavelength shift for BLI which will change after the formation of a complex. BLI has an additional similarity to our technology in that it doesn't use a flow cell to deliver antigen to an antibody, rather the antibody coated surface is introduced to a well containing antigen in solution. Despite this, BLI can regularly begin detecting antigen association almost immediately after the introduction of antigen[1,2] .This is primarily because many antibodies have been shown to have high association constants, a constant which describes that rate of complex formation as a function of the antigen and antibody's concentration. Using the association constant, we can predict the rate of complex formation for the conditions on our device. For example, the association constant for CR3022 and the SARs-CoV-2 spike protein has been estimated to be on the order of $1x10^6$ $M^{-1}s^{-1}$ [3]. We were able to observe complex formation when both the spike protein and HA were at approximately $100x10^{-21}$M and after coating with 25 µL of 50 µg/mL CR3022. Assuming complete capture of the antibody on the surface of the chip by PBASE (which was coated in molar excess relative to CR3022) we can calculate the theoretical maximum amount of CR3022 on the surface of the chip to be 1.25 µg.

$$Mass = Concentration\ x\ Volume$$

$$Mass = 50\frac{\mu g}{mL}\ x\ 0.025\ mL$$

$$Mass = 1.25\ \mu g$$

The molar mass of an IgG antibody is approximately 150,000 g/mol, thus a total of $8.33 \times 10^{-12}$ moles are present in each well coated with antibody. We can calculate the total number of antibody molecules on the surface by multiplying by Avogadro's number, a constant which describes the total number of molecular species per mole.

$$Number\ of\ antibodies = 8.33\ x\ 10^{-12}\ mol\ x\ 6.022\ x\ 10^{23} molecules/mol$$

$$Number\ of\ antibodies = 5.018\ x\ 10^{12}\ molecules$$

Looking at the "immune reaction" we can describe the formation of the rate complex by the following reaction below.

$$[CR3022][Spike] \underset{Kd}{\overset{Ka}{\longleftrightarrow}} [Complex]$$

Where [CR3022], [Spike], [Complex] represent the molar concentrations of CR3022, the Spike protein, and CR3022-Spike complexes respectively, and Ka and Kd are the association and dissociation constants. Expressing the rate of complex formation as a function of time we can write the rate of complex formation as follows:

$$\frac{d[Complex]}{dt} = K_a[CR3022][Spike] - K_d[Complex]$$

At time = 0, no complexes will have formed on the device yet. Additionally, the dissociation constant for CR3022 is several orders of magnitude lower than the association constant. Taking these two features into consideration we can estimate the initial rate of complex formation by the following equation.

$$\frac{d[Complex]}{dt} = K_a[CR3022][Spike]$$

The current form of this equation assumes that the CR3022 antibody is in solution, however all of the CR3022 is currently bound to the surface of the graphene. In this situation the concentration of CR3022 must be represented by its graphene surface density which can be simply calculated by dividing the total number of antibody molecules by the graphene surface area. For our device each well has a graphene surface area equal to 1.113 $mm^2$. This gives us a final surface area density of 4.5 x $10^{12}$ molecules/ $mm^2$

We now have all the information required to estimate the initial velocity of complex formation:

$$\frac{d[Complex]}{dt} = (1\text{x}10^6 M^{-1}s^{-1})\ \text{x}\ (4.5\ \text{x}\ 10^{12}\ \text{molecules/m}m^2)\ x\ (100x\ 10^{-21}M)$$

$$\frac{d[Complex]}{dt} = 0.45\frac{molecules}{mm^2 s}$$

By multiplying by the total area, we can predict that approximately 0.5 complexes can be formed every second. These calculations demonstrate that despite the extremely low concentrations of antigen, binding events are still expected to still occur in under 10 seconds. Under these ideal conditions, these calculations would suggest that the sensitivity of our device is approaching the order of magnitude of single molecule detection.

**Supplementary note 3**

The LOD values for breath biosensors[4] , mentioned in the main text, is the lowest possible limit of detection that may be required from a device for detecting the presence of the viral particles in a breath sample. The value has been calculated from the article which highlights the rough number of viral copies that may be available in a breath sample. Understanding that each viral copy weighs around 1fg[5]. For breath sample, the number of copies is around 10 copies/sample[4]. Within each COVID-19 particle or flu particle, there are ~23 spike or HA proteins on its surface [6]. Hence, this leads up ~23*$10^7$ spike proteins per sample setting the limit of detection for the breath sample at ~0.069 fg spike protein/sample, which is close to our limits.

Supplementary table 1

| Ref. | Ref (main text) | Method | | | | | Comments | Control | Antibody density measured |
|---|---|---|---|---|---|---|---|---|---|
| | | Sensing element | Linker | Probe | Charge screening strategy | Buffer concentration | | | |
| 7 | [50] | Reduced Graphene Oxide and Au nano particles | NHS-EDC | Phosphorodiamidate morpholino oligos | None | 0.01X PBS | 1) RGO has inferior carrier mobility. 2) NHS-EDC chemistry forms covalent bond, affecting sensitivity. 3) Non antibody probing agent, lower capture rates. 4) No functionalization for charge screening. | No | No |
| 8 | [23] | CVD Graphene | PBASE | COVID-19 Antibodies | None | Artificial saliva and UTM samples | No functionalisation for charge screening | No | No |
| 9 | [9] | Reduced Graphene Oxide and deposited Au | Cysteamine and NHS-EDC | COVID-19 Antibodies | None | 5.0 mM $[Fe(CN)_6]_3$ containing 0.1 M PBS and 0.1 M KCl | 1) RGO has inferior carrier mobility.2) NHS-EDC chemistry, covalent bond, affecting the sensitivity. 3) Non antibody probing agent, lower capture rates. 4) No functionalization for charge screening | No | No |
| 10 | [51] | PEDOT:PSS | 1,6-hexanedithiol (HDT) | COVID-19 nanobody | None | 1X PBS | 1) Oragnic transistors, reduced carrier mobility slow sensing. 2) No functionalisation for charge screening | No | Yes (Indirect measurement using QCM-D) |
| 11 | [52] | Graphite ink and magnetic beads | Adsorption based | COVID-19 Antibodies | None | DEA | 1) Graphite inks, inferior carrier mobility. 2) No charge screeining functionalisation. | No | No |
| 12 | [22] | CVD Graphene | PBASE | COVID-19 Antibodies | None | 1X pbs for testing and utm for clinical samples | No functionalisation for charge screening | No | No |
| 13 | [7] | Graphene Oxide and Au electrode | EDC: NHS | COVID-19 Antigen | None | 1X PBS | 1) RGO does not provide CVD graphene's carrier mobility. 2) NHS-EDC chemistry forms covalent bond, leading to sp3 hybridisation affecting the sensitivity of the device 3) No functionalisation for charge screening | No | No |
| 14 | [53] | PDPP-TT-based triblock copolymer (TBC) | Adsorption based | COVID-19 Antibody RBD | None | 1X PBS | 1) TBC prone to cracking, poor channel carrier ability. 2) No functionalization for charge screening. | No | No |
| 15 | [54] | CVD Graphene | PBASE | COVID-19 Antibodies | None | 1X PBS | No functionalisation for charge screening. | No | No |
| 16 | [55] | Carbon electrode with Cu2O nanocubes | ProtA | COVID-19 Antibodies | None | 1X PBS | 1) Screen printed carbon ink, inferior carrier mobility, low sensitivity.2) | No | No |

| | | | | | | | | | |
|---|---|---|---|---|---|---|---|---|---|
| | | | | | | | No functionalisation for charge screening. | | |
| 17 | [56] | SWCNTs | EDC: NHS | COVID-19 Antibodies | 4 % PEG | Nanopure water | NHS-EDC chemistry, covalent bond, affecting sensitivity. | No | No |
| 18 | [10] | $WSe_2$ | EDC: NHS | COVID-19 Antibodies | None | 0.01X PBS | 1) NHS-EDC chemistry, covalent bond, affecting sensitivity. 2) No functionalisation for charge screening | No | No |
| 19 | [13] | CVD Graphene | PBASE | Sialoglycan | None | 1X PBS | No functionalisation for charge screening | No | No |
| 20 | [15] | RGO Graphene | Immobili sation section of Long capture probe | Capture section of long capture probe | None | 1X PBS | 1) RGO has inferior carrier mobility. 2) Non antibody probing agent, lower capture rates. 3) No functionalization for charge screening | No | No |

(14) Ditte, K.; Anh, T.; Le, N.; Ditzer, O.; Sandoval Bojorquez, D. I.; Chae, S.; Bachmann, M.; Baraban, L.; Lissel, F. Rapid Detection of SARS-CoV-2 Antigens and Antibodies Using OFET Biosensors Based on a Soft and Stretchable Semiconducting Polymer. https://doi.org/10.1021/acsbiomaterials.1c00727.
